# Extracting Carotid Stenosis Severity from Clinical Notes Using Natural Language Processing: Development, Validation, and Application in a Nationwide Veteran Cohort

**DOI:** 10.1101/2025.05.16.25327773

**Authors:** Kyung Min Lee, Patrick R. Alba, Gina M. Biagetti, Anthony Gao, Ming Yin, Peter N. Danilov, Tia DiNatale, Cristina Perez, Katherine Hartmann, Gabrielle E. Shakt, Renae L. Judy, Tiffany R. Bellomo, Kathryn M. Pridgen, Philip S. Tsao, Naveen Balasundaram, Michael G. Levin, Scott M. Damrauer, Julie A. Lynch

**Affiliations:** VA Informatics and Computing Infrastructure (VINCI), VA Salt Lake City Health Care System, Salt Lake City, UT, USA; Division of Epidemiology, Department of Internal Medicine, University of Utah School of Medicine, Salt Lake City, UT, USA; Department of Surgery, Hospital of the University of Pennsylvania, Philadelphia, PA, USA; Department of Radiology, Hospital of the University of Pennsylvania, Philadelphia, PA, USA; Corporal Michael J. Crescenz VA Medical Center, Philadelphia, PA, USA; Department of Surgery, Perelman School of Medicine and the University of Pennsylvania, Philadelphia, PA, USA; Division of Vascular and Endovascular Surgery, Massachusetts General Hospital, Boston, MA, USA; Palo Alto Epidemiology Research and Information Center for Genomics, VA Palo Alto, CA, USA; Department of Medicine, Stanford University School of Medicine, Stanford, CA, USA; Department of Medicine, Perelman School of Medicine and the University of Pennsylvania, Philadelphia, PA, USA

## Abstract

**Importance:** Carotid stenosis, which is atherosclerotic narrowing of the extracranial carotid arteries, is an important risk factor for ischemic stroke. The prevalence of asymptomatic carotid stenosis is generally low, with moderate and severe stenosis present in up to ~6% and ~2% of the population, respectively. Prior studies of carotid stenosis have been small, and risk factors for carotid stenosis severity have been incompletely described. We sought to leverage the rich electronic health record data within the Veterans Health Administration to assess the prevalence and risk factors for carotid stenosis at the population level.

**Objective:** Develop and validate a natural language processing (NLP) tool to extract the ratio of peak systolic velocity of the internal carotid artery to that of the common carotid artery (ICA/CCA ratio) from carotid duplex ultrasound reports. Identify significant risk factors, presence and severity of carotid stenosis.

**Design:** Retrospective cross-sectional analysis

**Setting:** Veterans Health Administration (VHA) from 2001 to 2020

**Participants:** Veterans who underwent carotid artery duplex scans in the VHA from 2001 to 2020 and who had at least one valid ICA/CCA ratio. We excluded patients who had undergone carotid endarterectomy or stenting or who had a stroke or transient ischemic attack prior to the index date.

**Exposure(s):** Carotid artery duplex scan and cardiovascular disease risk factors including age, sex, self-identified race and ethnicity, healthcare utilization, smoking status, body mass index (BMI), blood pressure, hypertension, coronary heart disease, type 2 diabetes, and selected laboratory measures (i.e., hemoglobin A1c, low-density lipoprotein cholesterol, high-density lipoprotein cholesterol, triglyceride, and creatinine).

**Main outcome:** A categorical variable indicating carotid stenosis severity (<50%, 50-69%, ≥70%) based on ICA/CCA ratio (<2, ≥2 to <4, ≥4).

**Results:** The harmonic F1 score of the NLP tool was 0.907 for right value, 0.882 for left value, and 0.920 for max value. Among the 290,517 Veterans in the cohort, the median age was 68.2 years (IQR 61.9–75.0). 277,934 (95.7%) were males and 28,348 (10%) were of self-reported Black race. Black patients had 16% decreased risk of more severe carotid stenosis (OR 0.84, 95% CI 0.81–0.87, p<0.001). Yet, sensitivity analysis showed that among those with hemodynamically significant carotid stenosis (≥70% vs. 50–69%), Black race was associated with a 33% increased risk of severe carotid stenosis compared to White race (OR 1.33, 95% CI 1.23-1.43, p<0.001). All patient-level risk factors except high-density lipoprotein cholesterol were significantly associated with carotid stenosis severity.

**Conclusion and relevance:** The NLP tool performed well, and the study performed with our NLP-created cohort largely validates the risk factors identified by previous smaller studies, speaking to the validity of our tool to create usable cohorts for future large-scale research.

## INTRODUCTION

Carotid stenosis, which results from atherosclerotic plaque that causes a narrowing of the carotid artery, is a well-recognized risk factor for stroke, and the risk of stroke varies depending on the severity of stenosis.^1–3^ Asymptomatic carotid stenosis occurs in the absence of stroke or transient ischemic attack (TIA) history.^4^ The prevalence of asymptomatic carotid stenosis is generally low, with moderate and severe stenosis identified in up to ~6% and ~2% of the population, respectively.^5^ Carotid stenosis is typically diagnosed via carotid duplex ultrasound, a noninvasive means of measuring vessel diameter, visualizing atherosclerotic plaque, and capturing secondary signs of stenosis including changes in blood flow velocity. Diagnosis and risk stratification of carotid stenosis is made primarily using measures of the peak systolic velocity (PSV) of the internal carotid artery (ICA) and the common carotid artery (CCA) as well as the ratio of the former to the latter (ICA/CCA PSV ratio).^6,7^ Understanding carotid disease, particularly disease severity, is important because presence and severity of disease help guide intervention strategies.^8^

Our current knowledge of asymptomatic carotid stenosis comes mainly from relatively small cohorts,^3,9–11^ comprising only hundreds or thousands of participants. Using existing large datasets to create cohorts to study carotid stenosis severity has proved difficult because information about disease severity is generally included in patient notes and procedure reports as free text and not as structured data. As such, harnessing the power of natural language processing (NLP), which can be used to extract data from large electronic health records (EHR) datasets, may improve efficacy in epidemiologic research by making it easier to create larger, more diverse cohorts.^12^ Advances in NLP have enabled large-scale studies of EHR data such as radiology reports to identify individuals with carotid stenosis.^13–16^

In this study, we present the development and validation of an NLP tool to identify presence and severity of carotid stenosis within the Veteran’s Health Administration (VHA), the largest integrated healthcare system in the United States. This is the first NLP tool that extracts ICA/CCA ratio from carotid duplex ultrasound reports. Using the resulting cohort of Veterans, we performed the largest study of risk factors for asymptomatic carotid stenosis to date, demonstrating the utility of big data and NLP in carotid stenosis research.

## METHODS

We conducted a cross-sectional analysis of Veterans who received care at the VHA from 2001 to 2020. This study was approved by the institutional review boards of the VA Salt Lake City Healthcare System and the Corporal Michael J. Crescenz VA Medical Center.

### Data sources

Patient demographic and clinical data, including healthcare utilization, comorbidity, prescriptions, smoking status, body mass index (BMI), blood pressure, laboratory test results, and unstructured clinical notes containing reports of carotid artery ultrasounds were obtained from the Corporate Data Warehouse (CDW) of the VHA. The CDW is a national data repository that contains longitudinal clinical and administrative data for Veterans who received care at the VHA.

### Study sample

We identified active users of the VA from 2001 to 2020 who underwent a carotid artery duplex scan of either one or both carotid arteries, identified by current procedural terminology codes 93880 and 93882. We restricted the analysis to patients who had at least one valid ICA/CCA ratio, defined as a patient’s same-day duplex measurement with laterality being only one of the following: 1 left, 1 right, 1 left 1 right, 1 bilateral. We then identified the first valid duplex result for each patient. The greatest value of the first valid duplex result for each patient is the value used in analysis.

The index date represents the earliest date of all duplex scan results extracted by the NLP algorithm. We excluded patients who 1) had undergone carotid endarterectomy or stenting or 2) had a stroke or TIA prior to the index date.

### Development of NLP tool

We used the java-based LEO framework^17^ created by the VHA to develop and validate NLP tools that extracted and categorized duplex results based on whether these results were reported as quantitative, qualitative, or as the ICA/CCA ratio.

The initial NLP system was designed to identify four key concepts from carotid duplex studies; anatomy (internal carotid artery, discerning other non-relevant anatomies such as external carotid artery), laterality (left, right or bilateral), degree of stenosis expressed in qualitative terms or percentages (no stenosis, mild, moderate, severe, occluded, <50%, >50% etc.), and presence of ICA/CCA ratios.

Further development and validation of that NLP tool, which is reported here, focused on extracting ICA/CCA ratio results. The ICA/CCA ratio was extracted from the unstructured carotid artery duplex scan results using a rule-based NLP algorithm.

All NLP code for the system can be found at the following git repository: https://github.com/VINCI-AppliedNLP/CarotidStenosis.

#### Manual annotation

Annotation was completed by two nurse annotators, with the training batches double annotated and adjudicated by the study team. For development of the initial NLP tool, there were 100 distinct carotid duplex studies reviewed by two independent reviewers, with any discrepancies being adjudicated by a third, resulting in a Cohens Kappa Inter-annotator agreement of 0.83.

For the ICA/CCA ratio NLP tool, the annotation team documented the ICA/CCA ratios for the right and left carotid arteries from 300 carotid duplexes achieving an inter-annotator agreement with Cohen’s kappa greater than 0.85 across all batches annotated.

#### NLP validation

We assessed the performance of the NLP system using the following metrics: precision, recall (sensitivity), accuracy, and F-measure (F1 score) of the right value, the left value, and the maximum value found in a report. Performance was evaluated using the results of the 300 duplex scans that were manually annotated and held out from development. Ratios were rounded to the second decimal before being compared. For example, an annotated value 0.891 and an NLP extracted 0.89 would be counted as a true positive.

### Outcome

The outcome variable was a categorical variable indicating carotid stenosis severity as defined by ICA/CCA PSV ratio according to the currently recommended criteria for carotid interpretation (<2: normal/<50% stenosis; 2-4: 50–69% moderate stenosis; >4: ≥70% severe stenosis).^18,19^ Less than 50% stenosis is non-hemodynamically significant.

### Risk factors

The following patient-level risk factors were assessed at the index date (i.e., date of carotid duplex): age, sex, self-identified race and ethnicity, the total number of encounters (inpatient admissions or outpatient visits) in the prior two years, smoking status, BMI, systolic and diastolic blood pressures (SBP and DBP, respectively), indicator variables for pre-existing hypertension, coronary heart disease (CHD), and type 2 diabetes (T2D), and selected laboratory measures (i.e., hemoglobin A1c [HbA1c], low-density lipoprotein cholesterol [LDL-c], high-density lipoprotein cholesterol [HDL-c], triglyceride, and creatinine).

We obtained the most recent laboratory measures in the five years preceding the index date to minimize missing values. LDL-c and triglyceride levels were adjusted for lipid lowering therapies including statins and ezetimibe.^20^ We determined the receipt of prescriptions for selected drugs (statins, ezetimibe and other antilipemics, antiplatelets, anticoagulants, and acetylsalicylic acid; **Table B1**) in the 90 days prior to index date for descriptive analysis; however, we did not include them as risk factors in multivariable models due to the likely confounding by indication across the three categories of carotid stenosis severity. We included standardized BMI, blood pressures, and laboratory measures in all regression models to allow for comparisons of odds ratios of continuous variables with different units of measurement. We performed multiple imputation by predictive mean matching to impute missing BMI, blood pressures, and laboratory measures.^21,22^ The “ Other” racial category included American Indian or Alaska Natives, Native Hawaiian or Other Pacific Islanders, and Asian individuals.

### Statistical analysis

We performed Wilcoxon rank-sum tests for continuous variables and chi-squared tests for categorical variables to examine the differences in the included patient characteristics among carotid stenosis severity categories. We calculated bootstrap 95% confidence intervals of the differences in medians to compare the medians of continuous variables between two severity categories. We quantified the independent associations of patient-level risk factors on carotid stenosis severity by regressing carotid stenosis severity on each risk factor separately using ordinal logistic regression model.^23,24^ We estimated a multivariable ordinal logistic regression model to determine the associations of the risk factors with carotid stenosis severity as a three-level categorical variable. Ordinal logistic regression, used to model outcomes with ordered categories, assumes that the odds ratios across all categories are the same; biased estimates may result when this proportional odds assumption is violated.^25,26^ However, studies with large sample sizes such as the current analysis are less likely to violate the proportional odds assumption.^27,28^ Therefore, we chose ordinal logistic regression to model the currently recommended carotid stenosis severity categories in this analysis.

As a sensitivity analysis, we performed binary logistic regression analysis to make four pairwise comparisons of carotid stenosis severity categories: ≥50% vs. <50%, 50–69% vs. <50%, ≥70% vs. <50%, and ≥70% vs. 50–69%. By comparing ≥50% group (combination of 50–69% and ≥70% categories) with <50% category we examined the association of patient-level risk factors with the presence of hemodynamically significant stenosis (ICA/CCA PSV ratio>2). We then quantified the associations of risk factors with the presence of hemodynamically significant stenosis of varying severity (moderate or severe) by comparing <50% category with 50–69% and ≥70% categories separately. Finally, we compared ≥70% category with 50-69% category to investigate the associations of patient-level risk factors with having more severe stenosis. We performed the Z-test to compare odds ratios across different comparisons.^29^

We used SAS 9.4 to prepare the analytic dataset and R 4.4.1 to perform statistical tests and estimate regression models. All hypothesis tests were two-tailed with the significance level of 5%.

## RESULTS

We identified 13,058,116 Veterans who were active recipients of healthcare in the VHA between 2001 and 2020. There were 1,340,075 patients with at least one carotid duplex from 2001-2020, which included 2,594,346 distinct procedures. There were 974,889 (72.7%) patients who had 2,335,802 (90.3%) procedures with relevant radiology or clinical notes available for processing by the NLP tools. Of those, 414,455 patients had a report that included a valid ICA/CCA ratio value with laterality documented as left, right, or bilateral. There were 560,434 patients excluded whose reports did not include a valid ICA/CCA ratio. The majority were excluded because the ratio was missing from the report. In some cases, the ratio or laterality was ambiguous or not documented. Among the 414,455 Veterans with ICA/CCA ratio results, 290,517 had no carotid endarterectomy or stent procedures, and no stroke or TIA prior to the index date (**Figure 1**).

**Figure 1.**
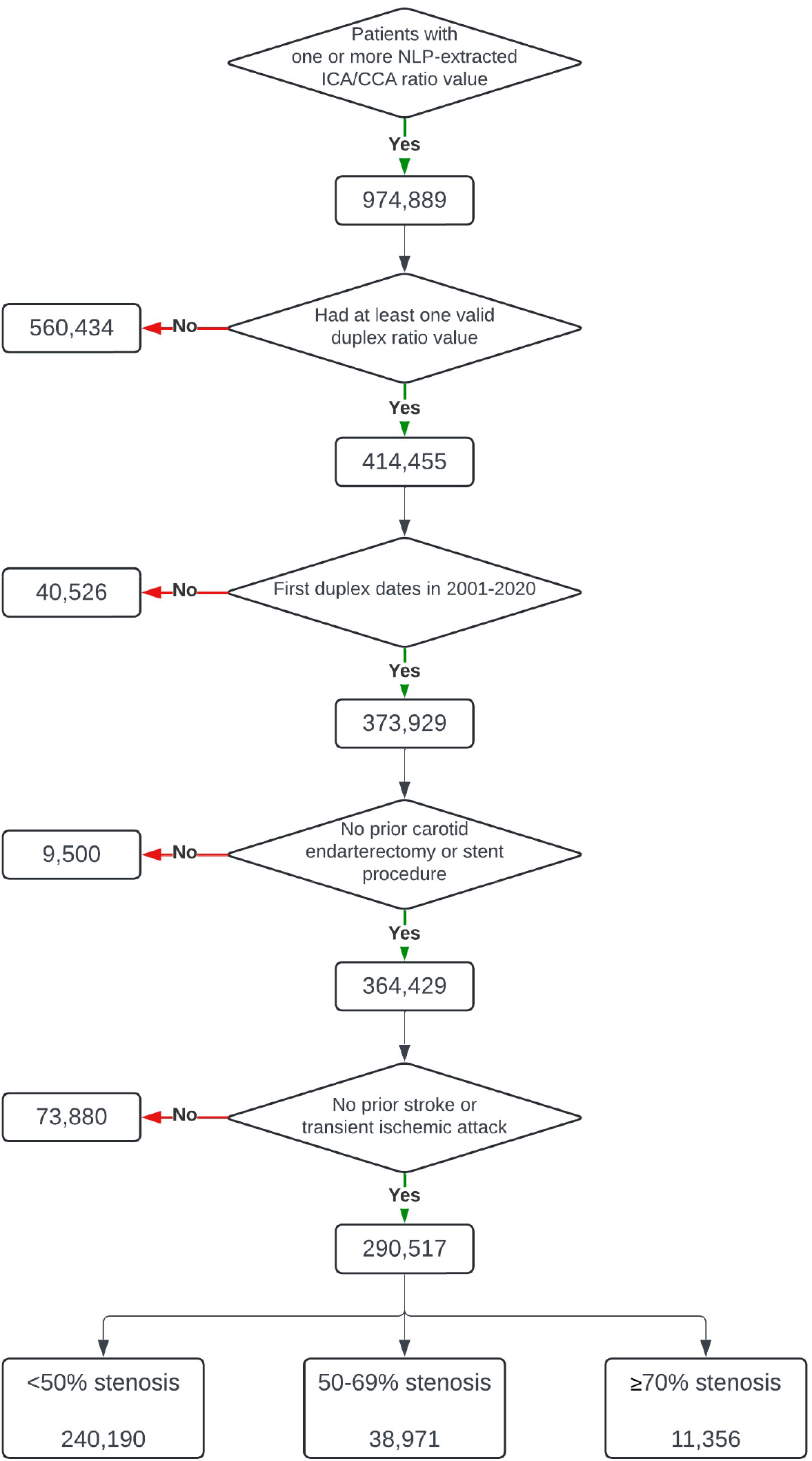
Study sample derivation

### NLP performance

Performance was evaluated against 300 held-out reports. Precision was >0.9 for all measurements (right value: 0.978; left value: 0.943; max value: 0.925), as was accuracy (right value: 0.940; left value: 0.927; max value: 0.943). Recall was 0.845 for right value, 0.828 for left value, and 0.916 for max value. F-measure was 0.907 for right value, 0.882 for left value, and 0.920 for max value. (**Table 1**) See Appendix A for additional information.

**Table 1.** NLP validation results.

| **Metric** | **Right Value Only** | **Left Value Only** | **Max Value Per Report** |
| --- | --- | --- | --- |
| True Positives (TP) | 88 | 82 | 98 |
| False Positives (FP) | 2 | 5 | 8 |
| True Negatives (TN) | 194 | 196 | 185 |
| False Negatives (FN) | 16 | 17 | 9 |
| Precision | 0.978 | 0.943 | 0.925 |
| Recall | 0.845 | 0.828 | 0.916 |
| Accuracy | 0.940 | 0.927 | 0.943 |
| F-Measure | 0.907 | 0.882 | 0.920 |

### Patient characteristics

Participants in the final study sample had a median age of 68.2 years (IQR 61.9–75.0). There were 277,934 (95.7%) men, 235,190 (81%) were of self-reported White race, and 28,348 (10%) were of self-reported Black race. The proportions of missing values for all imputed variables except HbA1c were less than 3% (**Table B2**). The distributions of the imputed variables remained unchanged after imputation.

Among the 290,517 Veterans with available duplex data and no symptoms of carotid disease 240,190 had ICA:CCA ratio <2, corresponding to a <50% stenosis, 38,971 had a ratio ≥2 and <4 corresponding to a 50% - 69% stenosis, and 11,356 had a ratio ≥4 corresponding to a ≥70% stenosis. Compared to those with <50% stenosis, patients with ≥70% stenosis were older (median difference 3.2 years, 95% CI 3.0–3.4) and more likely to have cardiovascular comorbidities (7%, 12%, and 3% higher rates of hypertension, CHD, and T2D, respectively; p<0.001). They were also more likely to be current smokers (28% vs. 23%, p<0.001), and had higher median levels of HbA1c (median difference 0.1%, 95% CI 0.1–0.1) and blood lipids except HDL-c (LDL-c: median difference 4.9 mg/dl, 95% CI 3.8–6.0; triglyceride: median difference 9.0 mg/dl, 95% CI 7.2–12.0; creatinine: median difference 0.1 mg/dl, 95% CI 0.1–0.1; **Table 2**).

**Table 2.** Patient characteristics.

|  | Overall | Carotid stenosis severity |  |  | p |
| --- | --- | --- | --- | --- | --- |
|  |  | <50% | 50-69% | ≥70% |  |
| **Total number of patients** | 290,517 | 240,190 | 38,971 | 11,356 |  |
| **Age (Q1-Q3)** | 68.2 (61.9-75.0) | 67.7 (61.4-74.3) | 70.6 (64.4-77.6) | 70.9 (65.1-77.9) | <0.001 |
| **Sex (%)** |  |  |  |  | <0.001 |
| Male | 277,934 (95.7) | 229,228 (95.4) | 37,638 (96.6) | 11,068 (97.5) |  |
| Female | 12,583 (4.3) | 10,962 (4.6) | 1,333 (3.4) | 288 (2.5) |  |
| **Race (%)** |  |  |  |  | <0.001 |
| White | 235,190 (81.0) | 193,250 (80.5) | 32,669 (83.8) | 9,271 (81.6) |  |
| Black | 28,348 (9.8) | 24,544 (10.2) | 2,799 (7.2) | 1,005 (8.8) |  |
| Other | 5,643 (1.9) | 4,919 (2.0) | 559 (1.4) | 165 (1.5) |  |
| Unknown | 21,336 (7.3) | 17,477 (7.3) | 2,944 (7.6) | 915 (8.1) |  |
| **Ethnicity (%)** |  |  |  |  | <0.001 |
| Not Hispanic or Latino | 267,991 (92.2) | 221,361 (92.2) | 36,118 (92.7) | 10,512 (92.6) |  |
| Hispanic or Latino | 11,572 (4.0) | 10,230 (4.3) | 1,062 (2.7) | 280 (2.5) |  |
| Unknown | 10,954 (3.8) | 8,599 (3.6) | 1,791 (4.6) | 564 (5.0) |  |
| **Healthcare utilization (Q1-Q3)** | 26.0 (14.0-46.0) | 26.0 (14.0-46.0) | 25.0 (13.0-45.0) | 24.0 (12.8-44.0) | <0.001 |
| **Comorbidities (%)** |  |  |  |  |  |
| Hypertension | 241,270 (83.0) | 196,693 (81.9) | 34,436 (88.4) | 10,141 (89.3) | <0.001 |
| Coronary heart disease | 114,185 (39.3) | 89,811 (37.4) | 18,820 (48.3) | 5,554 (48.9) | <0.001 |
| Type 2 diabetes | 114,422 (39.4) | 92,939 (38.7) | 16,685 (42.8) | 4,798 (42.3) | <0.001 |
| **Smoking Status (%)** |  |  |  |  | <0.001 |
| Never | 41,453 (14.3) | 36,131 (15.0) | 4,397 (11.3) | 925 (8.1) |  |
| Current | 68,987 (23.7) | 55,905 (23.3) | 9,870 (25.3) | 3,212 (28.3) |  |
| Former | 169,913 (58.5) | 139,913 (58.3) | 23,174 (59.5) | 6,826 (60.1) |  |
| Unknown | 10,164 (3.5) | 8,241 (3.4) | 1,530 (3.9) | 393 (3.5) |  |
| **Medication use (%)** |  |  |  |  |  |
| Statins | 170,900 (58.8) | 138,241 (57.6) | 25,110 (64.4) | 7,549 (66.5) | <0.001 |
| Other antilipemics | 23,969 (8.3) | 19,311 (8.0) | 3,700 (9.5) | 958 (8.4) | <0.001 |
| P2Y12 inhibitors | 25,606 (8.8) | 19,635 (8.2) | 4,544 (11.7) | 1,427 (12.6) | <0.001 |
| Other antiplatelets | 7,034 (2.4) | 5,353 (2.2) | 1,302 (3.3) | 379 (3.3) | <0.001 |
| Anticoagulants | 39,563 (13.6) | 31,857 (13.3) | 5,922 (15.2) | 1,784 (15.7) | <0.001 |
| Acetylsalicylic Acid | 70,154 (24.1) | 56,943 (23.7) | 10,069 (25.8) | 3,142 (27.7) |  |
| **Vitals** |  |  |  |  |  |
| *Body mass index* |  |  |  |  |  |
| Median (Q1-Q3) | 28.7 (25.4-32.5) | 28.8 (25.6-32.7) | 28.1 (24.9-31.8) | 27.7 (24.6-31.3) | <0.001 |
| Mean (SD) | 29.4 (5.8) | 29.5 (5.9) | 28.7 (5.6) | 28.3 (5.4) |  |
| *Systolic blood pressure (mmHg)* |  |  |  |  |  |
| Median (Q1-Q3) | 135.0 (122.0-148.0) | 134.0 (122.0-147.0) | 137.0 (124.0-151.0) | 139.0 (126.0-155.0) | <0.001 |
| Mean (SD) | 136.4 (20.2) | 135.8 (19.8) | 138.6 (21.8) | 141.3 (22.2) |  |
| *Diastolic blood pressure (mmHg)* |  |  |  |  |  |
| Median (Q1-Q3) | 76.0 (68.0-84.0) | 76.0 (69.0-84.0) | 74.0 (66.0-82.0) | 74.0 (66.0-82.0) | <0.001 |
| Mean (SD) | 76.0 (11.9) | 76.4 (11.8) | 74.1 (12.1) | 74.5 (12.1) |  |
| **Laboratory measures** |  |  |  |  |  |
| *HbA1c (%)* |  |  |  |  |  |
| Median (Q1-Q3) | 5.9 (5.6 - 6.6) | 5.9 (5.5 - 6.6) | 6.0 (5.6 - 6.8) | 6.0 (5.6 - 6.8) | <0.001 |
| Mean (SD) | 6.4 (1.6) | 6.4 (1.7) | 6.4 (1.6) | 6.5 (1.5) |  |
| *LDL-c (mg/dl)* |  |  |  |  |  |
| Median (Q1-Q3) | 132.7 (103.0 - 171.0) | 132.1 (103.0 - 170.7) | 134.2 (104.2 - 173.5) | 137.0 (106.0 - 178.1) | <0.001 |
| Mean (SD) | 144.2 (62.2) | 143.7 (62.0) | 145.6 (63.1) | 148.5 (64.4) |  |
| *HDL-c (mg/dl)* |  |  |  |  |  |
| Median (Q1-Q3) | 42.0 (35.0 - 52.0) | 42.0 (35.0 - 52.0) | 41.0 (35.0 - 51.0) | 42.0 (35.0 - 52.0) | <0.001 |
| Mean (SD) | 48.5 (26.0) | 48.5 (25.8) | 47.6 (25.4) | 50.5 (32.0) |  |
| *Triglyceride (mg/dl)* |  |  |  |  |  |
| Median (Q1-Q3) | 144.0 (97.8 - 217.0) | 143.0 (97.0 - 215.7) | 149.4 (101.3 - 224.0) | 152.0 (103.0 - 230.0) | <0.001 |
| Mean (SD) | 182.3 (170.0) | 180.7 (169.9) | 188.8 (170.0) | 193.0 (172.8) |  |
| *Creatinine (mg/dl)* |  |  |  |  |  |
| Median (Q1-Q3) | 1.0 (0.9 - 1.2) | 1.0 (0.9 - 1.2) | 1.1 (0.9 - 1.3) | 1.1 (0.9 - 1.3) | <0.001 |
| Mean (SD) | 6.5 (32.4) | 6.4 (31.9) | 7.0 (35.6) | 7.2 (31.6) |  |

Abbreviations: HbA1c, hemoglobin A1c; LDL-c, low-density lipoprotein cholesterol; HDL-c, high-density lipoprotein cholesterol

### Associations of patient-level risk factors with severity of carotid stenosis

All patient-level risk factors except HDL-c were significantly associated with carotid stenosis severity in a univariable analysis (**Table B3**). Older age, comorbidities, higher laboratory measures (except HDL-c), SBP, and current or former smoking were associated with an increased risk for more severe carotid stenosis.

The results of the multivariable ordinal logistic regression models are reported in **Table 3**. Every 10-year increase in age conferred a 22% increase in the likelihood of more severe carotid stenosis (odds ratio [OR] 1.022, 95% confidence interval [CI] 1.020–1.023, p<0.001). Compared to White patients, Black patients and those in the Other racial category had 16% and 26% decreased risk of more severe carotid stenosis, respectively (OR 0.84, 95% CI 0.81–0.87, p<0.001 for Black; OR 0.74, 95% CI 0.68-0.80 for Other). Individuals with pre-existing hypertension or CHD had a 44% increased risk of more severe carotid stenosis (OR 1.44, 95% CI 1.38–1.51 for hypertension; OR 1.44, 95% CI 1.41–1.47, p<0.001 for CHD). Each one standard deviation increase in HbA1c, LDL-c, HDL-c, triglyceride, and creatinine levels was associated with a 1–5% increased risk of more severe carotid stenosis. Current smokers were 67% more likely to experience more severe carotid stenosis compared with never smokers (OR 1.67, 95% CI 1.62–1.74, p<0.001).

**Table 3.** Association of patient-level factors with carotid stenosis severity from a multivariable ordinal logistic regression model.

|  | OR | 95% CI |  | p |
| --- | --- | --- | --- | --- |
| **Age at first duplex** | 1.02 | 1.02 | 1.02 | <0.001 |
| **Sex** |  |  |  |  |
| Male (Ref.) |  |  |  |  |
| Female | 1.02 | 0.96 | 1.07 | 0.553 |
| **Race** |  |  |  |  |
| White (Ref.) |  |  |  |  |
| Black | 0.84 | 0.81 | 0.87 | <0.001 |
| Other | 0.74 | 0.68 | 0.80 | <0.001 |
| Unknown | 0.90 | 0.86 | 0.94 | <0.001 |
| **Ethnicity** |  |  |  |  |
| Not Hispanic or Latino (Ref.) |  |  |  |  |
| Hispanic or Latino | 0.65 | 0.62 | 0.69 | <0.001 |
| Unknown | 1.19 | 1.13 | 1.26 | <0.001 |
| **Healthcare utilization** | 1.00 | 1.00 | 1.00 | <0.001 |
| **Comorbidities** |  |  |  |  |
| Hypertension | 1.44 | 1.38 | 1.51 | <0.001 |
| Coronary heart disease | 1.44 | 1.41 | 1.47 | <0.001 |
| Type 2 diabetes | 1.07 | 1.05 | 1.10 | <0.001 |
| **Laboratory measures** |  |  |  |  |
| HbA1c | 1.02 | 1.01 | 1.03 | <0.001 |
| LDL-c | 1.05 | 1.04 | 1.06 | <0.001 |
| HDL-c | 1.01 | 1.00 | 1.02 | 0.013 |
| Triglyceride | 1.06 | 1.05 | 1.07 | <0.001 |
| Creatinine | 1.02 | 1.01 | 1.02 | 0.001 |
| **Vitals** |  |  |  |  |
| Body mass index | 0.86 | 0.85 | 0.87 | <0.001 |
| Systolic blood pressure | 1.33 | 1.31 | 1.35 | <0.001 |
| Diastolic blood pressure | 0.76 | 0.75 | 0.77 | <0.001 |
| **Smoking status** |  |  |  |  |
| Never (Ref.) |  |  |  |  |
| Current | 1.67 | 1.62 | 1.74 | <0.001 |
| Former | 1.34 | 1.30 | 1.39 | <0.001 |
| Unknown | 1.37 | 1.29 | 1.45 | <0.001 |

Abbreviations: HbA1c, hemoglobin A1c; LDL-c, low-density lipoprotein cholesterol; HDL-c, high-density lipoprotein cholesterol

Our sensitivity analysis revealed that while the associations of patient-level risk factors remained practically unchanged when comparing patients with hemodynamically significant stenosis (≥50%) to those with non-hemodynamically significant stenosis (<50%), the associations of certain risk factors changed either in magnitude or in the direction of association in pairwise comparisons of carotid stenosis severity categories (**Table B4**). Among those with hemodynamically significant carotid stenosis (≥70% vs. 50–69%), Black race was associated with a 33% increased risk of severe carotid stenosis compared to White race (OR 1.33, 95% CI 1.23-1.43, p<0.001)..Pre-existing hypertension and CHD conferred smaller increased risks of severe carotid stenosis among patients with hemodynamically significant stenosis (hypertension: OR 1.18, 95% CI 1.05–1.32, p=0.005 in ≥70% vs. 50–69%, OR 1.68, 95% CI 1.52–1.87, p<0.001 in ≥70% vs. <50%, Z-test p<0.001; CHD: OR 1.08, 95% CI 1.03–1.13, p=0.002 in ≥70% vs. 50–69%, OR 1.51, 95% CI 1.45–1.58, p<0.001 in ≥70% vs. <50%, Z-test p<0.001). In ≥70% vs. <50% comparison, current smokers had more than two-fold increase in the risk of severe carotid stenosis (OR 2.37, 95% CI 2.20–2.56, p<0.001), which is markedly higher than the risk of increase in the other two comparisons (OR 1.51, 95% CI 1.45–1.57, p<0.001 in 50–69% vs. <50%, Z-test p<0.001; OR 1.55, 95% CI 1.43–1.69, p<0.001 in ≥70% vs. 50–69%, Z-test p<0.001).

## DISCUSSION

Our NLP algorithm identified 290,517 Veterans with ICA/CCA ratio values, resulting in a cohort >10-fold larger compared to previous studies.^13–16^ We then performed ordinal logistic regression analysis within this cohort to examine the association of known patient-level risk factors for carotid stenosis with severity of carotid stenosis.

The current study highlights the powerful potential of using NLP to mine data from emerging health record biobanks. This study leveraged EHR data to construct a cohort of patients screened for carotid stenosis using carotid duplex ultrasound and extract detailed phenotype data consisting of patients’ demographics, comorbidities, medication use, vitals, laboratory measures, and smoking status to facilitate the largest study to date of clinical risk factors for carotid stenosis. NLP algorithms have been used previously to extract documentation of a carotid stenosis diagnosis, however, to our knowledge this is the first study to extract more granular severity using ICA/CCA ratios. Further, the number of individuals in the current cohort is much larger compared to previous studies and may serve as a basis for further investigation in disease progression and outcomes.^13–16^

There have been previous limited attempts to extract carotid stenosis information from the EHR using NLP. In 2016, Mowery et al created an NLP system to identify reports with finding of significant carotid stenosis from radiology reports; however, this algorithm did not differentiate between moderate and severe stenosis.^14^ Our system builds up on this by now extracting quantitative measures of disease severity with high precision (right: 0.978; left: 0.943; max: 0.925) and accuracy (right: 0.940; left: 0.927; max: 0.943).

A different NLP system created by Chang et al (2021) does stratify by severity and also specifies the laterality of each case.^16^ This system identified normal carotid arteries with a precision of 0.98 and abnormal carotid arteries with a precision of 0.76, resulting in a cohort of over 15,000 patients and demonstrating the potential utility of NLP to create large cohorts. Our system, when applied to the VHA EHR, was able to generate a dataset approximately 20 times larger than that of Chang et al.

An error analysis of the system performance when compared to the held-out data set revealed the most common errors emanating from random variation in the text format that was not accounted for in the training data. While values were accurately captured from many templates and semi-structured forms, these errors primarily resulted from ad hoc templates and document structures created within the note. The majority of false positives occurred due to clinical guidelines presented in the note being misinterpreted as test results. Despite this the overall false positive rate was low (2.2% to 7.6% depending on measure and laterality).

Using our NLP extracted data we identified a cohort of 290,517 asymptomatic Veterans with carotid duplex measurements for analysis. We found that older age, pre-existing hypertension, CHD, and T2D, higher levels of HbA1c, LDL-c, HDL-c, triglyceride, creatinine, higher SBP, and smoking was associated with higher likelihood of experiencing severe carotid stenosis, largely validating the risk factors identified in previous, smaller studies.^10,11,30,31^ The fact that the study done within our NLP-created cohort validates previous studies speaks to the legitimacy of our tool to create usable cohorts for large-scale research.

Our findings indicated that the well-established risk factors for atherosclerotic cardiovascular disease (ASCVD) are also significant risk factors for not only the likelihood of having a carotid stenosis but also the likelihood of having a more severe carotid stenosis. The results of this study suggest that the general approaches to primary and secondary prevention of ASCVD should also be effective in preventing carotid stenosis across the disease severity spectrum. Except for age, sex, and race/ethnicity, many of the traditional atherosclerotic risk factors including dyslipidemia, hypertension, diabetes, and smoking are modifiable through lifestyle and behavioral changes. The largest odds ratios were seen with smoking, hypertension, and CHD, suggesting that management of these modifiable risk factors through established prevention strategies^32^ may decrease severity of carotid stenosis.

The associations of self-identified race and ethnicity with carotid stenosis severity suggests that individuals of non-White racial groups and Hispanic patients were less likely to experience severe carotid stenosis compared to White and non-Hispanic patients, respectively. The lower odds of having a hemodynamically significant carotid stenosis among Black patients and Hispanic patients compared to White patients and non-Hispanic patients, respectively, are consistent with previous studies that reported lower prevalence of carotid stenosis among Black patients and Hispanic patients.^33–36^ However, results of the sensitivity analysis suggest that of those with hemodynamically significant stenosis (i.e., ≥50%), Black patients were more likely to present with severe rather than moderate stenosis compared to White patients.

These seemingly paradoxical findings may be attributable to several reasons. First, index event bias is potentially present as Black patients may be less likely to receive carotid imaging.^37^ Since the current study analyzed individuals who had received a clinical duplex scan, Black patients may have been disproportionately excluded from the study due to their lower likelihood of receiving a scan.^26–28^ Additionally, the Black patients’ higher likelihood of experiencing a more severe carotid stenosis among a subset of patients with hemodynamically significant stenosis may be due to treatment disparity as Black patients may be less likely to undergo carotid endarterectomy^38,39^ and more likely to experience delays in receiving treatment.^40^ The weaker associations of age and comorbidities seen among those with hemodynamically significant stenosis in the sensitivity analysis are likely due to survivorship bias, as older patients are more likely to have died before developing severe carotid stenosis (≥70%).

This study has other limitations as well. The VHA’s EHR data are subject to the limitations of any other large-scale EHR data including missing data, potential inconsistencies in coding practices across different medical centers and unreliability of self-reported race and ethnicity. We attempted to minimize the influence of missing data by implementing multiple imputation by predictive mean matching for continuous variables and including “ Unknown” categories for categorical variables. Despite the promising performance in identifying ICA/CCA ratios, our NLP system still exhibits some limitations, primarily related to occasional variations in report text formats that might cause false negatives or missed literalities. While racially and ethnically diverse, the Veteran patient population in the VHA consists of predominantly older men, thereby limiting the generalizability of the study results.

## CONCLUSIONS

The rules-based NLP system reliably identified degree of stenosis in our patient population, allowing for the largest study of clinical risk factors for carotid stenosis to date. The risk factors we identified for both the presence and severity of carotid stenosis largely validated previous studies on the subject, suggesting that this NLP tool may be able to create large cohorts for future research studies. The risk factors were also similar to well-established risk factors ASCVD, suggesting that prevention strategies used for CVD may also reduce the risk of carotid stenosis across the disease spectrum.

## Data Availability

Deidentified patient-level data are currently accessible to all VA researchers with appropriate IRB approvals.

## ACKNOWLEDGEMENTS

This research was supported by the Department of Veterans Affairs, Veterans Health Administration, Office of Research and Development, Biomedical Laboratory Research and Development Service Award I01-BX0061459 and IK2-BX006551. This work was supported using resources and facilities of the Department of Veterans Affairs (VA) Informatics and Computing Infrastructure (VINCI), including data analytics conducted by its Precision Medicine research team, which is funded under the research priority to Put VA Data to Work for Veterans (VA ORD 24-D4V-02). The views expressed in this article are those of the authors and do not necessarily reflect the position or policy of the Department of Veterans Affairs or the United States government.

JAL, PRA, PND, TD, and KMP report grants from Alnylam Pharmaceuticals, Inc., AstraZeneca Pharmaceuticals LP, Biodesix, Inc., Janssen Pharmaceuticals, Inc., Novartis International AG, Parexel International Corporation through the University of Utah or Western Institute for Veteran Research outside the submitted work. MGL reports grants from the Doris Duke Foundation (2023-0224, research funding to the institution from MyOme, and consulting fees from BridgeBio outside the submitted work. SMD reports grants from the National Heart Lung and Blood Institute, in kind support from Novo Nordisk, and consulting fees from Tourmaline Bio, outside the current work.

