## Appendix A for "Extracting Carotid Stenosis Severity from Clinical Notes Using Natural Language Processing: Development, Validation, and Application in a Nationwide Veteran Cohort"

### Appendix A. Additional description of natural language processing algorithm used to extract ICA/CCA PSV ratios

#### 1. Introduction

The goal was to develop an NLP algorithm to extract the ICC/CCA PSV ratios found in reports from carotid artery ultrasounds. The system was developed using the java-based LEO framework developed at the United States Department of veterans Affairs. These are the results of the system performance when compared to a held-out set of annotated data. All NLP code for this system can be found at the following git repository:

https://github.com/VINCI-AppliedNLP/CarotidStenosis.

#### 2. Evaluation Metrics

**Precision:** The accuracy of positive predictions. It is calculated as:
Precision = TP / (TP + FP)

**Recall (Sensitivity):** The ability to identify all positive samples. It is calculated as:
Recall = TP / (TP + FN)

**Accuracy:** The fraction of correct predictions. It is calculated as:
Accuracy = (TP + TN) / (TP + TN + FP + FN)

**F-Measure (F1 Score):** The harmonic mean of precision and recall. It is given by:
F1 Score = 2 x (Precision x Recall) / (Precision + Recall)

#### 3. Detailed Error Analysis

Some of the challenges and discrepancies observed in the results include:

1. **False Negatives**: There were a few instances, especially on the left side, where the NLP system missed ratios present in the reports. These errors were all due to random variation in the text format that was not accounted for in the training data. While values were accurately captured from many templates and semi-structured forms, the primarily cause of these errors were a result of ad hoc templates and document structures created within the note. E.g.
   "ICA/CCA Ratio:
   Right: 1
   LEFT: 1.6"
2. **Extracted Value, Missed laterality**. It should be noted, however, that when the left or right laterality was missed, a ratio value was frequently still extracted, just without the laterality being correctly attributed. Of the 16 FN Right values, 6 were correctly extracted without laterality and would be present if only the max laterality per report is used. Of the 17 FN Left values, this is true for 3.

3. **False Positives**: The system exhibited a relatively low rate of false positives. Of the false positives that were observed, the majority occurred when a clinical guideline was used to describe ICC/CCA ratios and was not identified by the system. i.e. the following example being extracted as (Ratio = 2)
   “ICA/CCA Peak Systolic Ratios:
   < 2 correlates with < 50% stenosis“

**Figure A1. Distribution of values found in the annotation reports**

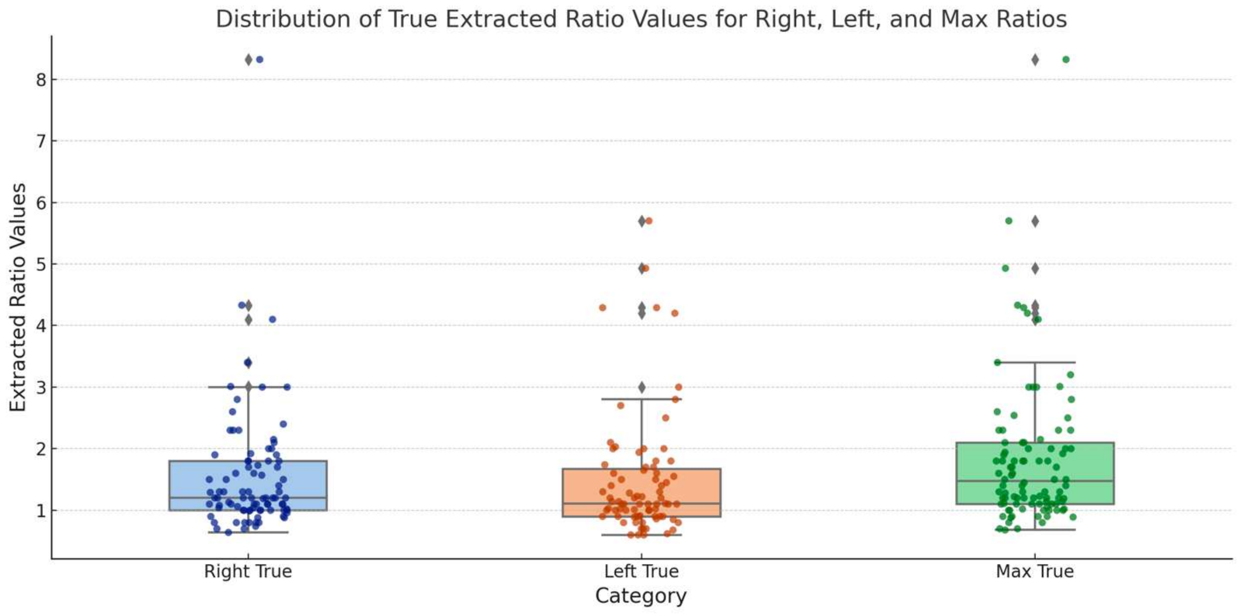

**Figure A2. Distribution of values when extracted by NLP, split further by the FP, and TP cases. This figure shows that infrequent random errors by NLP still fell within the same ranges of extracted correct values.**

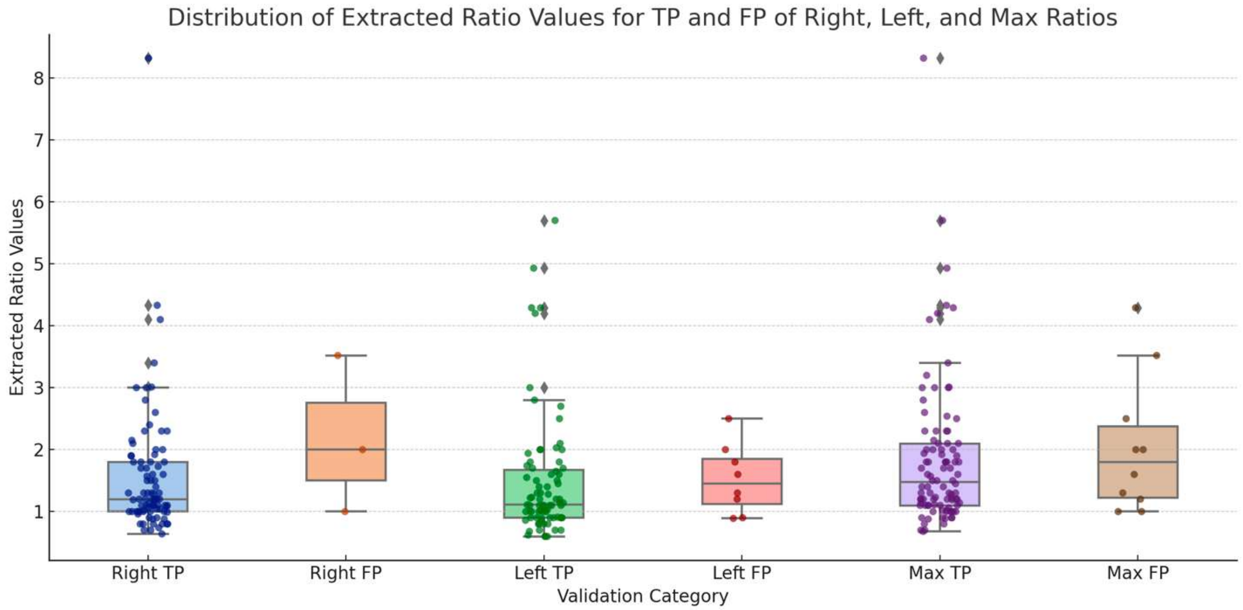
