## Appendix B for "Extracting Carotid Stenosis Severity from Clinical Notes Using Natural Language Processing: Development, Validation, and Application in a Nationwide Veteran Cohort"

### Appendix B. Supplementary tables

**Table B1**. List of selected medications
**Table B2**. Summary of missing data and multiple imputation
**Table B3**. Independent associations of patient-level factors with carotid stenosis severity from univariable ordinal logistic regression models
**Table B4**. Pairwise comparisons of carotid stenosis severity categories

**Table B1**. **List of selected medications**

| Drug group | Drug |
|---|---|
| Statins | Atorvastatin |
| | Fluvastatin |
| | Lovastatin |
| | Pitavastatin |
| | Pravastatin |
| | Rosuvastatin |
| | Simvastatin |
| Other antilipemics | Ezetimibe |
| | Alirocumab |
| | Evolocumab |
| | Bempedoic Acid |
| | Evinacumab |
| | Lomitapide |
| | Fenofibrate |
| | Gemfibrozil |
| | Bezafibrate |
| | Cholestyramine |
| | Colesevelam |
| | Colestipol |
| | Niacin |
| P2Y12 inhibitors | Clopidogrel |
| | Prasugrel |
| | Ticagrelor |
| | Cangrelor |
| Other antiplatelets | Abciximab |
| | Eptifibatide |
| | Tirofiban |
| | Dipyridamole |
| | Cilostazol |
| | Ticlopidine |
| Anticoagulants | Heparin |
| | Warfarin |
| | Rivaroxaban |
| | Dabigatran |
| | Apixaban |
| | Edoxaban |
| | Enoxaparin |
| | Fondaparinux |
| Acetylsalicylic Acid | Aspirin |

**Table B2. Summary of missing data and multiple imputation**

| Variable | Observations | Number of missing values | % missing | Mean | SD | Min | Max |
|---|---|---|---|---|---|---|---|
| **Body mass index** | | | | | | | |
| Before | 289,751 | 766 | 0.26% | 0.0 | 1.0 | -3.9 | 36.6 |
| After | 290,517 | | | 0.0 | 1.0 | -3.9 | 36.6 |
| **Systolic blood pressure** | | | | | | | |
| Before | 290,134 | 383 | 0.13% | 0.0 | 1.0 | -5.9 | 48.3 |
| After | 290,517 | | | 0.0 | 1.0 | -5.9 | 48.3 |
| **Diastolic blood pressure** | | | | | | | |
| Before | 290,081 | 436 | 0.15% | 0.0 | 1.0 | -6.4 | 10.7 |
| After | 290,517 | | | 0.0 | 1.0 | -6.4 | 10.7 |
| **HbA1c** | | | | | | | |
| Before | 229,427 | 61,090 | 21.03% | 0.0 | 1.0 | -3.8 | 54.0 |
| After | 290,517 | | | -0.1 | 1.0 | -3.8 | 54.0 |
| **LDL-c** | | | | | | | |
| Before | 282,645 | 7,872 | 2.71% | 0.0 | 1.0 | -2.3 | 24.4 |
| After | 290,517 | | | 0.0 | 1.0 | -2.3 | 24.4 |
| **HDL-c** | | | | | | | |
| Before | 283,349 | 7,168 | 2.47% | 0.0 | 1.0 | -1.8 | 28.2 |
| After | 290,517 | | | 0.0 | 1.0 | -1.8 | 28.2 |
| **Triglyceride** | | | | | | | |
| Before | 283,509 | 7,008 | 2.41% | 0.0 | 1.0 | -1.1 | 72.9 |
| After | 290,517 | | | 0.0 | 1.0 | -1.1 | 72.9 |
| **Creatinine** | | | | | | | |
| Before | 285,942 | 4,575 | 1.57% | 0.0 | 1.0 | -0.2 | 127.3 |
| After | 290,517 | | | 0.0 | 1.0 | -0.2 | 127.3 |

Abbreviations: HbA1c, hemoglobin A1c; LDL-c, low-density lipoprotein cholesterol; HDL-c, high-density lipoprotein cholesterol

**Table B3. Independent associations of patient-level factors with carotid stenosis severity from univariable ordinal logistic regression models**

| | OR | 95% CI | | p |
|---|---|---|---|---|
| **Age at first duplex** | 1.03 | 1.03 | 1.03 | <0.001 |
| **Sex** | | | | |
| Male (Ref.) | | | | |
| Female | 0.69 | 0.66 | 0.73 | <0.001 |
| **Race** | | | | |
| White (Ref.) | | | | |
| Black | 0.72 | 0.69 | 0.75 | <0.001 |
| Other | 0.68 | 0.63 | 0.74 | <0.001 |
| Unknown | 1.02 | 0.98 | 1.06 | 0.265 |
| **Ethnicity** | | | | |
| Not Hispanic (Ref.) | | | | |
| Hispanic or Latino | 0.62 | 0.59 | 0.66 | <0.001 |
| Unknown | 1.30 | 1.24 | 1.36 | <0.001 |
| **Healthcare utilization** | 1.00 | 1.00 | 1.00 | <0.001 |
| **Comorbidities** | | | | |
| Hypertension | 1.97 | 1.88 | 2.06 | <0.001 |
| Coronary heart disease | 1.66 | 1.63 | 1.69 | <0.001 |
| Type 2 diabetes | 1.12 | 1.10 | 1.15 | <0.001 |
| **Laboratory measures** | | | | |
| HbA1c | 1.05 | 1.04 | 1.06 | <0.001 |
| LDL-c | 1.04 | 1.03 | 1.05 | <0.001 |
| HDL-c | 0.99 | 0.98 | 1.00 | 0.124 |
| Triglyceride | 1.05 | 1.04 | 1.06 | <0.001 |
| Creatinine | 1.02 | 1.01 | 1.03 | <0.001 |
| **Vitals** | | | | |
| Body mass index | 0.84 | 0.84 | 0.85 | <0.001 |
| Systolic blood pressure | 1.18 | 1.17 | 1.19 | <0.001 |
| Diastolic blood pressure | 0.83 | 0.82 | 0.83 | <0.001 |
| **Smoking status** | | | | |
| Never (Ref.) | | | | |
| Current | 1.60 | 1.55 | 1.66 | <0.001 |
| Former | 1.47 | 1.42 | 1.51 | <0.001 |
| Unknown | 1.59 | 1.50 | 1.68 | <0.001 |

Abbreviations: HbA1c, hemoglobin A1c; LDL-c, low-density lipoprotein cholesterol; HDL-c, high-density lipoprotein cholesterol

**Table B4**. **Pairwise comparisons of carotid stenosis severity categories**

| | **≥50% (vs. <50%/Normal)** | | | | **50-69% (vs. <50%/Normal)** | | | | **≥70 (vs. <50%/Normal)** | | | | **≥70% (vs. 50-69%)** | | | |
|---|---|---|---|---|---|---|---|---|---|---|---|---|---|---|---|---|
| | **OR** | **95% CI** | | **p** | **OR** | **95% CI** | | **p** | **OR** | **95% CI** | | **p** | **OR** | **95% CI** | | **p** |
| **Age at first duplex** | 1.02 | 1.02 | 1.02 | <0.001 | 1.02 | 1.02 | 1.02 | <0.001 | 1.03 | 1.02 | 1.03 | <0.001 | 1.01 | 1.00 | 1.01 | <0.001 |
| **Sex** | | | | | | | | | | | | | | | | |
| Male (Ref.) | | | | | | | | | | | | | | | | |
| Female | 1.03 | 0.97 | 1.09 | 0.332 | 1.08 | 1.02 | 1.15 | 0.011 | 0.82 | 0.73 | 0.93 | 0.002 | 0.77 | 0.67 | 0.88 | <0.001 |
| **Race** | | | | | | | | | | | | | | | | |
| White (Ref.) | | | | | | | | | | | | | | | | |
| Black | 0.83 | 0.80 | 0.86 | <0.001 | 0.78 | 0.75 | 0.81 | <0.001 | 0.99 | 0.93 | 1.06 | 0.787 | 1.33 | 1.23 | 1.43 | <0.001 |
| Other | 0.73 | 0.68 | 0.79 | <0.001 | 0.72 | 0.66 | 0.79 | <0.001 | 0.77 | 0.66 | 0.90 | 0.001 | 1.06 | 0.89 | 1.26 | 0.516 |
| Unknown | 0.90 | 0.86 | 0.94 | <0.001 | 0.88 | 0.84 | 0.93 | <0.001 | 0.95 | 0.87 | 1.03 | 0.209 | 1.07 | 0.97 | 1.17 | 0.158 |
| **Ethnicity** | | | | | | | | | | | | | | | | |
| Not Hispanic (Ref.) | | | | | | | | | | | | | | | | |
| Hispanic or Latino | 0.65 | 0.62 | 0.69 | <0.001 | 0.66 | 0.62 | 0.71 | <0.001 | 0.61 | 0.54 | 0.69 | <0.001 | 0.92 | 0.81 | 1.06 | 0.237 |
| Unknown | 1.19 | 1.13 | 1.26 | <0.001 | 1.19 | 1.12 | 1.27 | <0.001 | 1.21 | 1.09 | 1.35 | <0.001 | 1.03 | 0.92 | 1.16 | 0.576 |
| **Healthcare utilization** | 1.00 | 1.00 | 1.00 | <0.001 | 1.00 | 1.00 | 1.00 | <0.001 | 1.00 | 1.00 | 1.00 | <0.001 | 1.00 | 1.00 | 1.00 | 0.008 |
| **Comorbidities** | | | | | | | | | | | | | | | | |
| Hypertension | 1.44 | 1.37 | 1.51 | <0.001 | 1.39 | 1.32 | 1.47 | <0.001 | 1.68 | 1.52 | 1.87 | <0.001 | 1.18 | 1.05 | 1.32 | 0.005 |
| Coronary heart disease | 1.44 | 1.41 | 1.47 | <0.001 | 1.41 | 1.38 | 1.45 | <0.001 | 1.51 | 1.45 | 1.58 | <0.001 | 1.08 | 1.03 | 1.13 | 0.002 |
| Type 2 diabetes | 1.07 | 1.05 | 1.10 | <0.001 | 1.08 | 1.06 | 1.11 | <0.001 | 1.05 | 1.00 | 1.09 | 0.045 | 0.96 | 0.91 | 1.01 | 0.110 |
| **Laboratory measures** | | | | | | | | | | | | | | | | |
| HbA1c | 1.02 | 1.01 | 1.03 | 0.001 | 1.02 | 1.00 | 1.03 | 0.011 | 1.03 | 1.01 | 1.05 | 0.001 | 1.03 | 1.00 | 1.06 | 0.055 |
| LDL-c | 1.04 | 1.03 | 1.05 | <0.001 | 1.04 | 1.02 | 1.05 | <0.001 | 1.07 | 1.05 | 1.09 | <0.001 | 1.03 | 1.01 | 1.06 | 0.002 |
| HDL-c | 1.01 | 1.00 | 1.02 | 0.128 | 0.98 | 0.97 | 0.99 | 0.005 | 1.08 | 1.06 | 1.10 | <0.001 | 1.09 | 1.07 | 1.11 | <0.001 |
| Triglyceride | 1.06 | 1.05 | 1.07 | <0.001 | 1.05 | 1.04 | 1.06 | <0.001 | 1.07 | 1.05 | 1.08 | <0.001 | 1.04 | 1.02 | 1.06 | <0.001 |
| Creatinine | 1.02 | 1.01 | 1.03 | 0.001 | 1.01 | 1.00 | 1.02 | 0.008 | 1.02 | 1.00 | 1.04 | 0.016 | 1.00 | 0.98 | 1.02 | 0.731 |
| **Vitals** | | | | | | | | | | | | | | | | |
| Body mass index | 0.86 | 0.85 | 0.87 | <0.001 | 0.88 | 0.86 | 0.89 | <0.001 | 0.82 | 0.80 | 0.84 | <0.001 | 0.94 | 0.92 | 0.96 | 0.000 |
| Systolic blood pressure | 1.33 | 1.31 | 1.34 | <0.001 | 1.29 | 1.27 | 1.30 | <0.001 | 1.46 | 1.43 | 1.49 | <0.001 | 1.13 | 1.10 | 1.16 | <0.001 |
| Diastolic blood pressure | 0.76 | 0.75 | 0.77 | <0.001 | 0.76 | 0.75 | 0.78 | <0.001 | 0.73 | 0.72 | 0.75 | <0.001 | 0.96 | 0.94 | 0.99 | 0.003 |
| **Smoking status** | | | | | | | | | | | | | | | | |
| Never (Ref.) | | | | | | | | | | | | | | | | |
| Current | 1.66 | 1.60 | 1.72 | <0.001 | 1.51 | 1.45 | 1.57 | <0.001 | 2.37 | 2.20 | 2.56 | <0.001 | 1.55 | 1.43 | 1.69 | <0.001 |
| Former | 1.33 | 1.29 | 1.38 | <0.001 | 1.25 | 1.21 | 1.29 | <0.001 | 1.75 | 1.63 | 1.87 | <0.001 | 1.38 | 1.28 | 1.50 | <0.001 |
| Unknown | 1.37 | 1.29 | 1.45 | <0.001 | 1.33 | 1.24 | 1.41 | <0.001 | 1.58 | 1.40 | 1.79 | <0.001 | 1.19 | 1.04 | 1.36 | 0.011 |

Abbreviations: HbA1c, hemoglobin A1c; LDL-c, low-density lipoprotein cholesterol; HDL-c, high-density lipoprotein cholesterol
